# Persistent SARS-CoV-2 presence is companied with defects in adaptive immune system in non-severe COVID-19 patients

**DOI:** 10.1101/2020.03.26.20044768

**Authors:** Bing Liu, Junyan Han, Xiaohuan Cheng, Long Yu, Li Zhang, Wei Wang, Lan Ni, Chaojie Wei, Yafei Huang, Zhenshun Cheng

**Author notes:** **Corresponding author:** Zhenshun Cheng MD; Department of Respiratory and Critical Care Medicine, Zhongnan Hospital of Wuhan University, Wuhan, China. **Alternate corresponding author:** Yafei Huang MD, PhD, Department of Pathogen Biology, School of Basic Medicine, Tongji Medical College, Huazhong University of Science and Technology, Wuhan, China. Author Y.H. and Z.C. contributed equally to this manuscript.

## Abstract

**Background:** COVID-19 has been widely spreading. We aim to examine adaptive immune cells in non-severe patients with persistent SARS-CoV-2 shedding.

**Methods:** 37 non-severe patients with persistent SARS-CoV-2 presence transferred to Zhongnan hospital of Wuhan University were retrospectively recruited to PP (persistently positive) group, which was further allocated to PPP group (n=19) and PPN group (n=18), according to their testing results after 7 days (N=negative). Epidemiological, demographic, clinical and laboratory data were collected and analyzed. Data from age- and sex-matched non-severe patients at disease onset (PA [positive on admission] patients, n=37), and lymphocyte subpopulation measurements from matched 54 healthy subjects were extracted for comparison.

**Results:** Compared with PA patients, PP patients had much improved laboratory findings, including WBCs, neutrophils, lymphocytes, neutrophil-to-lymphocyte ratio, albumin, AST, CRP, SAA, and IL-6. The absolute numbers of CD3^+^ T cells, CD4^+^ T cells, and NK cells were significantly higher in PP group than that in PA group, and were comparable to that in healthy controls. PPP subgroup had markedly reduced B cells and T cells compared to PPN group and healthy subjects. Finally, paired results of these lymphocyte subpopulations from 10 PPN patients demonstrated that the number of T cells and B cells significantly increased when the SARS-CoV-2 tests turned negative.

**Conclusion:** Persistent SARS-CoV-2 presence in non-severe COVID-19 patients is associated with reduced numbers of adaptive immune cells. Monitoring lymphocyte subpopulations could be clinically meaningful in identifying fully recovered COVID-19 patients.

**Summary:** Defects in adaptive immune system, including reduced T cells and B cells, were frequently observed in non-severe COVID-19 patients with persistent SARS-CoV-2 shedding. Assessment of immune system could be clinically relevant for discharge management.

## Introduction

Emerged in Wuhan, China in December 2019, COVID-19 has quickly spread to other provinces of China, and to 170 other countries and areas across six continents. As of March 24, 372,757 cases and 16,231 deaths has been confirmed, making COVID-19 a formidable threat to global health [1]. Despite the high mortality rate of 4.2% worldwide, around 85% of patients infected with SAR-CoV-2 developed non-severe disease, as recently reported by two large epidemiological studies [2,3]. These patients appeared to have a better clinical outcome compared to those with severe disease [2]. However, the accumulating cases, together with limited availability of hospital beds still resulted in a run on medical resources in China and several other countries.

Upon supportive and anti-viral treatments, most of the patients with non-severe disease achieved clinical cure, as demonstrated by attenuated symptoms, as well as improved laboratory and imaging findings [2,4]. However, pathogenic cure in terms of viral RNA-conversion, had to be reached before patients being discharged from hospitals for the purpose of disease control, at least in China. The kinetics of viral shedding [6,7], the possible presence of viral RNA in multiple sites [8-11], sampling error and the technical limitations of RT-PCR sometimes led to a false testing result [12,13], posing a great challenge to discharge management after obtaining clinical cure. Nevertheless, a recent investigation reported that the median duration of viral shedding was 20 days after disease onset for patients infected with SARS-CoV-2 [4]. The fact that almost half of the patients remained positive for viral RNA 20 days post disease onset, together with the aforementioned limitations of viral RNA detection method, suggest that other indicators should be used in combination with viral RNA test to achieve a better discharge management for non-severe COVID-19 patients, and to save limited hospital beds for those severe patients.

Changes in lymphocyte compartment have been reported to be correlated with disease course, illness severity and clinical outcome [14-17]. However, measurement of lymphocyte subpopulations has yet to be investigated in non-severe COVID-19 patients with persistent viral RNA presence. This study was therefore designed to examine the correlation of changes in adaptive immune cells with persistent viral RNA presence in non-severe COVID-19 patients, and to evaluate its potential application in discharge management for these patients.

## Methods

### Participants, study design and definitions

This study was approved by Zhongnan Hospital of Wuhan University (ZHWU)’s ethical review board (No. 2020013). All hospitalized patients admitted to ZHWU from January 16 to February 27, 2020 with confirmed COVID-19 was included for initial screen. Written informed consent was waived by the Ethics Commission of the hospital for emerging infectious diseases.

Disease status was defined according to the guideline of SARS-CoV-2 (Trial Version 5 of the Chinese National Health Commission) [5]: (i) mild, with slight clinical symptoms but no imaging presentations of pneumonia; (ii) moderate, with fever, respiratory tract and other symptoms, and imaging findings of pneumonia; (iii) severe, with any of the following conditions: respiratory distress, respiratory frequency ≥30 times/minutes, finger oxygen saturation at rest ≤93%, or oxygenation index [PaO2/FiO2]≤300 mmHg (1 mmHg=0·133 kPa); (iiii) critical, with any of the following conditions: respiratory failure requires mechanical, ventilation, shock, combined with other organ failure requires intensive care unit care and treatment. Patients with mild or moderate illness was considered to have non-severe disease. Disease status was monitored up to March 6, 2020, the final date of follow-up.

Non-severe COVID-19 patients that were tested SARS-CoV-2 positive for more than 20 days after diagnosis were retrospectively allocated to the PP group. Age- and sex-matched healthy subjects and non-severe COVID-19 patients were randomly recruited into the HC group and the PA group, respectively.

### Data collection

Demographic information, clinical characteristics (including medical history, exposure history, comorbidities, surgery history, signs, and symptoms), chest computed tomographic (CT) scan or X-ray results, and laboratory findings of each patient were obtained from the electronic medical record system of ZHWU and analyzed by three independent researchers.

### Laboratory testing

Patient nasopharyngeal swab specimens were collected for the SARS-CoV-2 viral nucleic acid detection using real-time reverse transcriptase-polymerase chain reaction (RT-PCR) assay. The viral nucleic acid testing for all patients was performed by the clinical laboratory from Zhongnan Hospital of Wuhan University in Wuhan. Detailed protocol was described previously [8]. Lymphocyte subpopulations were examined by FACS Aria III cytometer (BD bioscience, USA) and analyzed using Flowjo software v.10.2 (BD bioscience, USA). Other laboratory indicators, including blood routine, C-reactive protein (CRP), serum amyloid A (SAA), and IL-6, were collected for each patients.

### Statistical Analysis

Data analysis was performed using SPSS (Statistical Package for the Social Sciences, version 23). Categorical variables were reported as absolute (relative frequencies) and compared by χ^2^ tests or Fisher’s exact tests. Continuous variables were expressed as mean (SD) if they are normally distributed or median (interquartile range, IQR) if they are not and compared by independent group t tests or Mann-Whitney U tests, respectively. *P*<0·05 was considered as statistically significant.

## Results

### Baseline characteristics

After initial screen, 37 non-severe COVID-19 patients that were tested positive for SARS-CoV-2 more than 20 days were recruited to the PP group. The median age for these patients was 53 years (IQR 45-60; Table 1), and 25 (67.6%) patients were men. Since no patients had direct exposure history of Huanan seafood market, we presumed all patients in this study were community-infected cases. The most common symptoms at onset of illness were fever (78.4%) and dry cough (78.4%), followed by dyspnea (29.7%), expectoration (24.3%), and diarrhea (13.5%). The less common symptoms included pharyngalgia (2.7%), hemoptysis (2.7%) and weep tears (2.7%). Common complications included CVD (13.5%), followed by diabetes (5.4%) and hepatitis (5.4%). There were 3 current smokers. The baseline characteristics were summarized in Table 1.

**Table 1.** Baseline Characteristics

| **Median**<br>**(IQR)** | **HC (n=54)** | **COVID-19 Patients** |  |  |  | **P Value** |  |  |
| --- | --- | --- | --- | --- | --- | --- | --- | --- |
|  |  | **PA (n=37)** | **PP (n=37)** | **PPP (n=19)** | **PPN (n=18)** | **PP vs HC** | **PP vs PA** | **PPP vs PPN** |
| **Age** | 56 (39-65) | 54 (42-66) | 53 (45-60) | 50 (44-58) | 55 (47-62) | 0.37 | 0.25 | 0.33 |
| **Sex** |  |  |  |  |  |  |  |  |
| Female | 21 (38.9) | 14 (37.8.) | 12(32.4) | 3(15.8) | 9(50.0) | 0.52 | 0.62 | **0.02** |
| Male | 33 (61.1) | 23 (62.2) | 25(67.6) | 16(84.2) | 9(50.0) | 0.52 | 0.62 | **0.02** |
| **Symptoms** |  |  |  |  |  |  |  |  |
| Fever |  |  | 29 (78.4) | 16 (84.2) | 13 (74.2) |  |  | 0.38 |
| Cough |  |  | 29 (78.4) | 17(89.5) | 12(66.7) |  |  | 0.09 |
| Expectoration |  |  | 9(24.3) | 7(16.8) | 2(11.1) |  |  | 0.07 |
| Hemoptysis |  |  | 1 (2.7) | 1 (5.3) | 0 (0) |  |  | 0.32 |
| Dyspnea |  |  | 11 (29.7) | 3 (15.8) | 8 (44.4) |  |  | 0.06 |
| Weep tears |  |  | 1 (2.7) | 1 (5.3) | 0 (0) |  |  | 0.32 |
| Pharyngalgia |  |  | 1 (2.7) | 0 (0) | 1 (5.3) |  |  | 0.30 |
| Diarrhea |  |  | 5 (13.5) | 3 (15.8) | 2 (11.1) |  |  | 0.68 |
| **Comorbidities** |  |  |  |  |  |  |  |  |
| CVD |  |  | 5 (13.5) | 3 (15.8) | 2 (11.1) |  |  | 0.68 |
| Diabetes |  |  | 2 (5.4) | 2 (10.5) | 0 (0) |  |  | 0.16 |
| Hepatitis |  |  | 2 (5.4) | 1 (5.3) | 1 (5.6) |  |  | 1 |
| Other |  |  | 1 (2.7) | 0 (0) | 1 (5.6) |  |  | 0.30 |
| **Treatments** |  |  |  |  |  |  |  |  |
| GC |  |  | 2 (5.4) | 0 (0) | 2 (11.1) |  |  | 0.14 |
| Antiviral |  |  | 26 (70.3) | 15 (78.9) | 11 (61.1) |  |  | 0.23 |
| Antibiotics |  |  | 17 (45.9) | 7 (36.8) | 10 (55.6) |  |  | 0.52 |
| TCM |  |  | 30 (81.1) | 17 (89.5) | 13 (72.2) |  |  | 0.18 |
| **Others** |  |  |  |  |  |  |  |  |
| Smoke |  |  | 3 (8.1) | 0 (0) | 3 (16.7) |  |  | 0.06 |
| Drink |  |  | 3 (8.1) | 0 (0) | 3 (16.7) |  |  | 0.06 |

Data are median (IQR) or n (%). *P* values were obtained from $\chi^2$ tests, Fisher's exact tests, T tests or Mann–Whitney U tests, when appropriate. *P* < 0.05 was considered statistically significant (in bold).
*Abbreviations:* HC, healthy controls; COVID-19, coronavirus disease 19; PA (positive on admission); PP (persistently positive); PPP, PP patients tested positive again; PPN: PP patients tested negative; CVD: cardiovascular diseases; GC, glucocorticoids; TCM, traditional Chinese medicine.

### Blood cell counts, blood biochemicals and inflammatory biomarkers in patients with COVID-19

Table 2 presented the laboratory testing results of these patients (PP group) on admission to our hospital. Unfortunately, the results of the same patients at disease onset were not available since these patients were first admitted to mobile cabin hospital and then transferred to our hospital, we therefore randomly selected another 37 age- and sex-matched COVID-19 patients confirmed with non-severe disease (PA group), who had their blood test at disease onset on admission to our hospital, for comparison. Compared with patients from the PA group, those from the PP group had significantly higher numbers of lymphocytes (1.5 [1.3-1.8] *vs* 0.9 [0.7-1.3] ×10^9^/L; p<0.001) and higher concentrations of ALB (42.5 [41.7-43.7] *vs* 39.3 [37.7-41.3] g/L; p=0.02), but much lower NLR (1.8 [1.5-2.4] *vs* 2.7 [1.7-4.9]; p=0.01), as well as lower levels of CRP (1.8 [0.9-2.6] *vs* 10.8 [2.7-36.7] g/L; p<0.001), SAA (6.4 [4.5-10.5] *vs* 48.4 [16.1-96.7] mg/L; p<0.001), and IL-6 (2.3 [1.5-2.9] *vs* 6.2 [1.8-16.6] mg/L; p<0.001). The differences were even more pronounced upon using reference ranges to determine the abnormalities, patients from the PP group had much less frequent abnormal results for WBC (5.4% *vs* 40.5%; p<0.001), neutrophils (10.8% *vs* 43.2%; p=0.002), lymphocytes (13.5% *vs* 72.8%; p<0.001), PLTs (8.1% *vs* 35.1%; p=0.005), CRP (5.4% *vs* 43.2%; p<0.001), SAA (27.0% *vs* 56.8%; p=0.01) and IL-6 (2.7% *vs* 48.6%; p<0.001), as compared to those from the PA group. In together, these results demonstrated that PP patients, upon treatment in mobile cabin hospital and transferred to our hospital, had much improved laboratory findings than PA patients at disease onset, even though they had persistent SARS-CoV-2 shedding.

**Table 2.** Laboratory results of COVID-19 patients.

| Median (IQR) | Normal range | PA (n=37) | PP (n=37) | P Value |
| --- | --- | --- | --- | --- |
| **Blood cells** |  |  |  |  |
| WBC (x10<sup>9</sup>/L) | 3.5-9.5 | 3.9 (3.1-56.4) | 4.98 (4.4-5.7) | 0.45 |
| Abnormal No. (%) |  | 15 (40.5) | 2 (5.4) | **<0.001** |
| Neutrophils (x10<sup>9</sup>/L) | 1.8-6.3 | 2.4 (1.6-4.1) | 2.9 (2.3-3.5) | 0.53 |
| Abnormal No. (%) |  | 16 (43.2) | 4 (10.8) | **0.002** |
| Lymphocytes (x10<sup>9</sup>/L) | 1.1-3.2 | 0.9 (0.7-1.3) | 1.5 (1.3-1.8) | **<0.001** |
| Abnormal No. (%) |  | 27 (72.8) | 5 (13.5) | **<0.001** |
| NLR |  | 2.7 (1.7-4.9) | 1.8 (1.5-2.4) | **0.01** |
| PLTs (x10<sup>9</sup>/L) | 125-350 | 181 (127-201) | 182 (155-220) | 0.85 |
| Abnormal No. (%) |  | 13 (35.1) | 3 (8.1) | **0.005** |
| **Blood biochemicals** |  |  |  |  |
| Hb (g/L) | 130-175 | 132 (111-139) | 132 (123-145) | 0.43 |
| Abnormal No. (%) |  | 16 (43.2) | 10 (27.0) | 0.14 |
| ALB (g/L) | 40-55 | 39.3 (37.7-41.3) | 42.5 (41.7-43.7) | **0.02** |
| Abnormal No. (%) |  | 19 (51.3) | 11 (29.7) | 0.06 |
| ALT (U/L) | 9-50 | 21.0 (18.3-34.5) | 27.0 (20.5-42.0) | 0.12 |
| Abnormal No. (%) |  | 3 (8.1) | 6 (16.2) | 0.29 |
| AST (U/L) | 15-40 | 21.5 (17.0-26.0) | 25.0 (19.0-32.0) | 0.81 |
| Abnormal No. (%) |  | 4 (10.8) | 10 (27.0) | 0.08 |
| Total Bilirubin (μmol/L) | 5-21 | 11.2 (8.6-13.4) | 12.9 (10.9-16.8) | 0.35 |
| Abnormal No. (%) |  | 3 (8.1) | 2 (5.4) | 0.64 |
| **Inflammatory biomarkers** |  |  |  |  |
| CRP (g/L) | 0-10 | 10.8 (2.7-36.7) | 1.8 (0.9-2.6) | **<0.001** |
| Abnormal No. (%) |  | 16 (43.2) | 2 (5.4) | **<0.001** |
| SAA (mg/L) | 0-10 | 48.4 (16.1-96.7) | 6.4 (4.5-10.5) | **<0.001** |
| Abnormal No. (%) |  | 21 (56.8) | 10 (27.0) | **0.01** |
| IL-6 (pg/mL) | 0-7 | 6.2 (1.8-16.6) | 2.3 (1.5-2.9) | **<0.001** |
| Abnormal No. (%) |  | 18 (48.6) | 1 (2.7) | **<0.001** |

Data are median (IQR) or n (%). *P* values were obtained from $\chi^2$ tests, Fisher's exact tests, T tests or Mann-Whitney U tests, when appropriate. *P* < 0.05 was considered statistically significant (in bold).
*Abbreviations:* COVID-19, coronavirus disease 19; PA (positive on admission); PP (persistently positive); **NLR**: Neutrophil-to-lymphocyte ratio; PLTs, platelets; Hb, hemoglobin; ALB, albumin; ALT, alanine aminotransferase; AST, aspartate aminotransferase; CRP, C-reactive protein; SAA, serum amyloid A.

### Lymphocyte subsets in peripheral blood

It has been reported that dysregulated immune response were correlated with the severity of COVID-19 [16]. However, changes in adaptive immune cells in non-severe COVID-19 patients with persistent SARS-CoV-2 shedding has yet to be examined. For this purpose, peripheral blood samples from patients in the PA and PP group were collected, the absolute numbers and relative frequencies of each lymphocyte subpopulations were compared between these two groups. In addition, 54 age- and sex-matched healthy subjects were randomly selected as healthy control (the HC group). As shown in Table 3, we failed to find any differences between the PP group and the HC group, but patients from both groups had increased numbers of CD3^+^ T cells, CD4^+^ T cells, and NK cells compared to those from the PA group. In addition, PA patients had significantly lower frequency of B cells compared with healthy subjects (Table 3). These results indicated that non-severe COVID-19 patients (PA group) have already dysregulated immune system at diasese onset, and those with persistent SARS-CoV-2 shedding could restore this abnormality to some level.

**Table 3.** Lymphocyte subpopulations in periphery blood of COVID-19 patients and healthy controls.

| Median (IQR) | Normal range | HC (n=54) | PA (n=37) | PP (n=37) | P Value |  |  |
| --- | --- | --- | --- | --- | --- | --- | --- |
|  |  |  |  |  | PP vs PA | PP vs HC | PA vs HC |
| Absolute numbers /µl |  |  |  |  |  |  |  |
| CD3<sup>+</sup> T cells | 955.0-2860.0 | 1091.0 | 683.0 | 1083.0 | **0.02** | 0.39 | **0.006** |
| CD4<sup>+</sup> T cells | 550.0-1440.0 | 655.5 | 420.0 | 611.0 | **0.04** | 0.94 | **0.013** |
| CD8<sup>+</sup> T cells | 320.0-1250.0 | 345 .5 | 276.5 | 382.0 | 0.068 | 0.337 | 0.09 |
| B cells | 240.0-560.0 | 171 .0 | 176.0 | 162.0 | 0.81 | 0.92 | 0.86 |
| NK cells | 150.0-1100.0 | 276.5 | 149.5 | 243.0 | **0.03** | 0.74 | **0.001** |
| Frequencies of lymphocytes (%) |  |  |  |  |  |  |  |
| CD4<sup>+</sup> T cells | 27.0-51.0 | 39.1 (33.5-46.3) | 38.4 (33.3-44.5) | 41.2 (32.6-44.7) | 0.46 | 0.59 | 0.79 |
| CD8<sup>+</sup> T cells | 15.0-44.0 | 23.5 (18.4-29.3) | 25.4 (21.6-33.2) | 26.0 (22.3-31.2) | 0.59 | 0.76 | 0.49 |
| CD4<sup>+</sup>/ CD8<sup>+</sup> T cells | 0.9-2.0 | 1.8 (1.1-2.3) | 1.3 (1.0-1.8) | 1.5 (1.2-1.9) | 0.95 | 0.17 | 0.17 |
| B cells | 5.0-18.0 | 11.6 (7.2-15.8) | 15.7 (8.8-19.9) | 12.6 (8.3-17.4) | 0.08 | 0.61 | **0.02** |
| NK cells | 7.0-40.0 | 17.3 (11.7-24.8) | 14.9 (8.9-21.3) | 14.1 (12.5-20.8) | 0.40 | 0.60 | 0.79 |

Data are median (IQR) or n (%). *P* values were obtained from $\chi^2$ tests , Fisher's exact tests, T tests or Mann–Whitney U tests, when appropriate. *P* < 0.05 was considered statistically significant (in bold).
*Abbreviations:* COVID-19, coronavirus disease 19; PA (positive on admission); PP (persistently positive); NK, natural killer.

Upon admission, PP patients received the same standard treatment in our hospital. After at least 7 days, 18 of them that were tested negative for SARS-CoV-2 in two consecutive examinations were retrospectively allocated to the PPN group, and 19 of them who remained positive at the same time point were designated as PPP patients. Of note, the PPP group have more males than the PPN group (86.4% [16 of 19] vs 50% [9 of 18]; p=0.02; Table 1). We did not find any differences in symptoms and laboratory findings for these two groups (supplementary Table 1 and 2). However, when lymphocyte subpopulations were examined, PPP patients were found to have significantly lower numbers of CD3^+^ T cells (p=0.001), CD4^+^ T cells (p=0.005), CD8^+^ T cells (p=0.003), and B cells (p=0.005), but higher proportion of NK cells (p=0.02) than PPN patients (Fig 1A and 1B). Next, we determined the abnormalities for each parameters by using reference ranges published elsewhere (Table 3, Fig 2A and 2B) [16]. Similar trends were found in CD3^+^ T cells (p=0.001), CD4^+^ T cells (p=0.001), CD8^+^ T cells (p=0.01), and B cells (p<0.001). Since the reference ranges of lymphocyte subpopulations were established based on all Chinese Han population, we therefore selected 54 age- and sex-matched healthy subjects from Wuhan for comparison. Again, PPP patients exhibited much less numbers of CD3^+^ T cells (p=0.044), CD4^+^ T cells (p=0.034), and B cells (p=0.02) than healthy subjects (Fig 2C and 2D).

**Figure 1.**
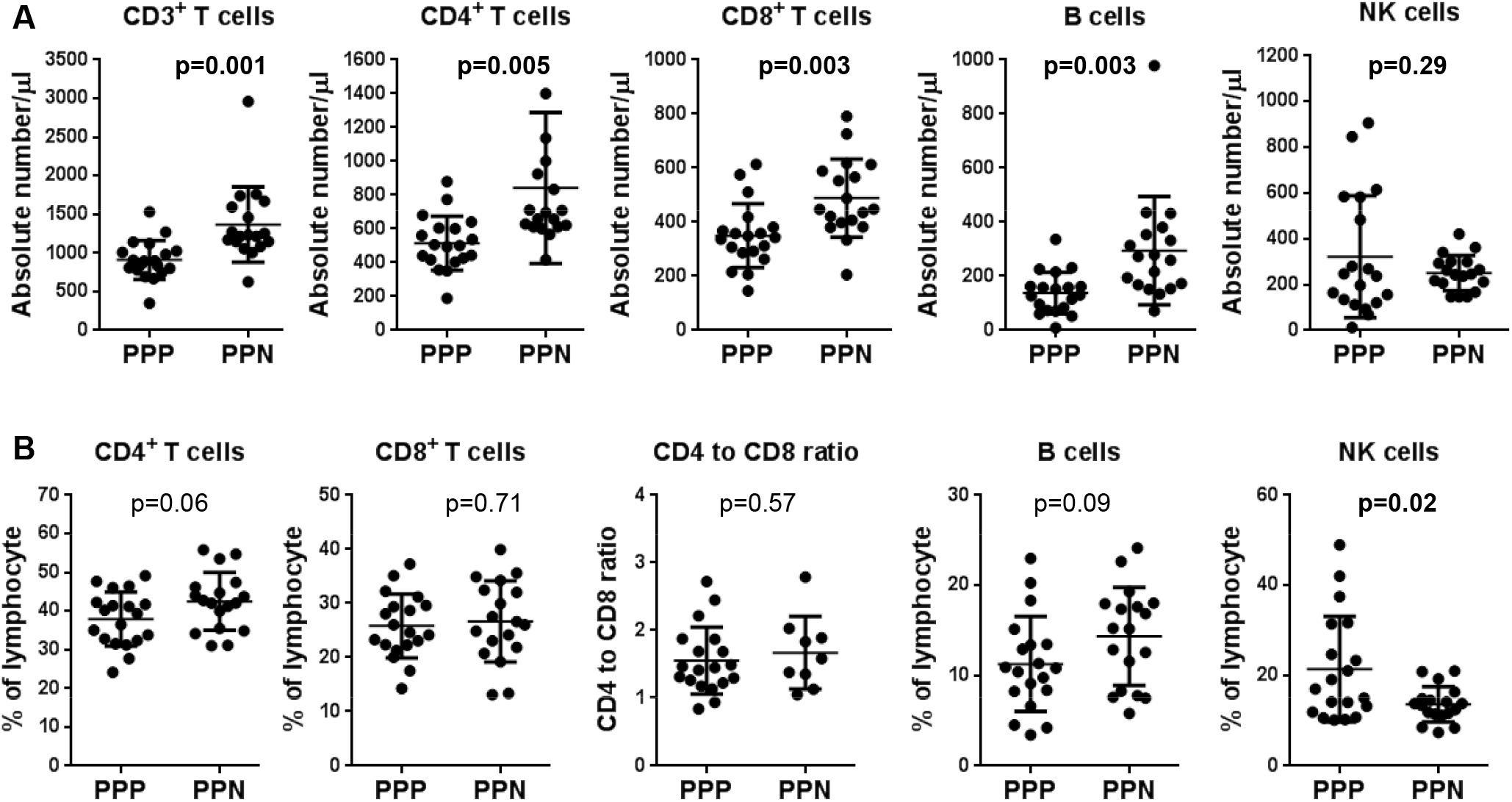
Absolute numbers (A) and relative frequencies of lymphocyte subpopulations (B) in peripheral blood of PP patients were tested positive again at least 7 days after they were admitted to our hospital (PPP), and PP patients were tested negative in 7 days after they were admitted to our hospital (PPN). *P* < 0.05 was considered statistically significant (in bold).

**Figure 2.**
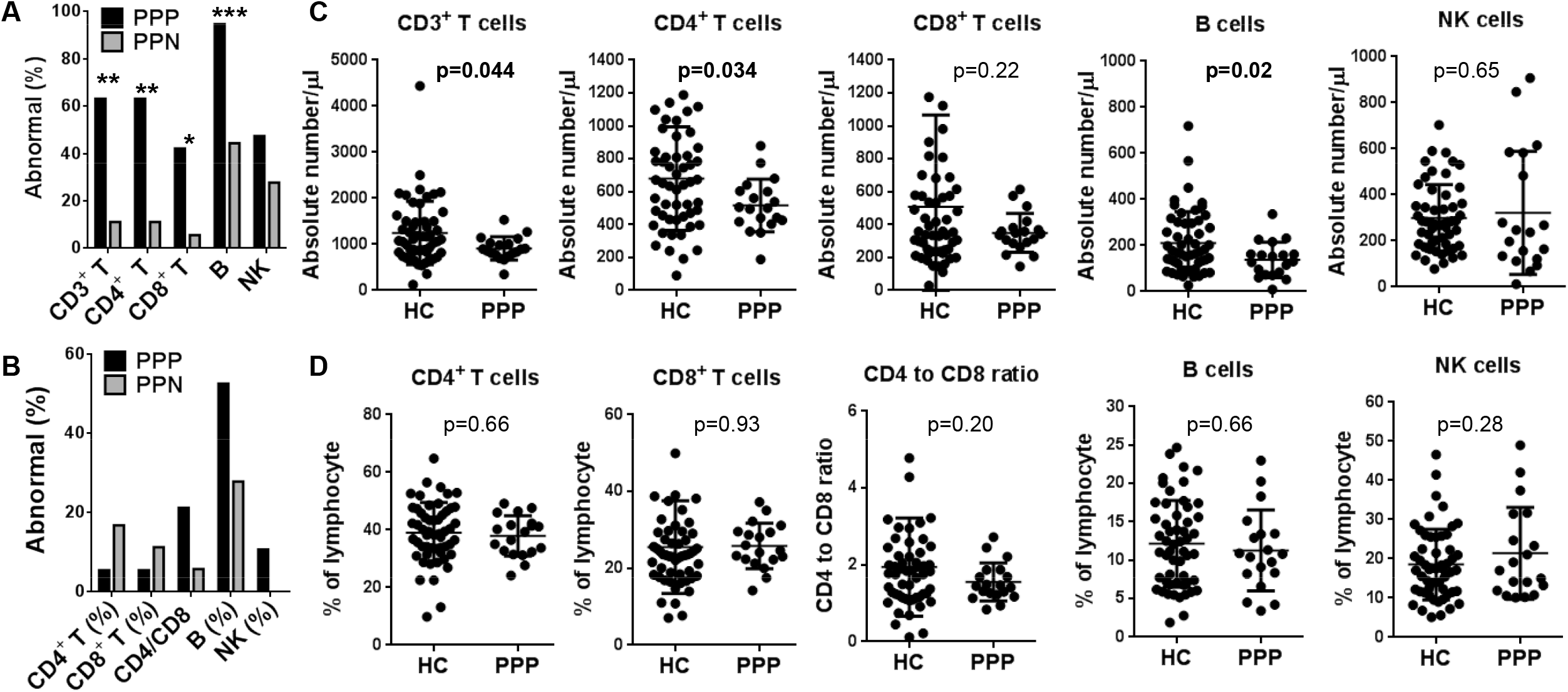
The proportion of abnormalities of lymphocyte subpopulations in terms of absolute numbers (A) and relative frequencies of (B) in peripheral blood of PPP and PPN patients, and absolute numbers (C) and relative frequencies of lymphocyte subpopulations (D) in peripheral blood of PPP patients and healthy controls (HC). * *P* < 0.05; ** *P* < 0.01; *** *P* < 0.001.

**Figure 3.**
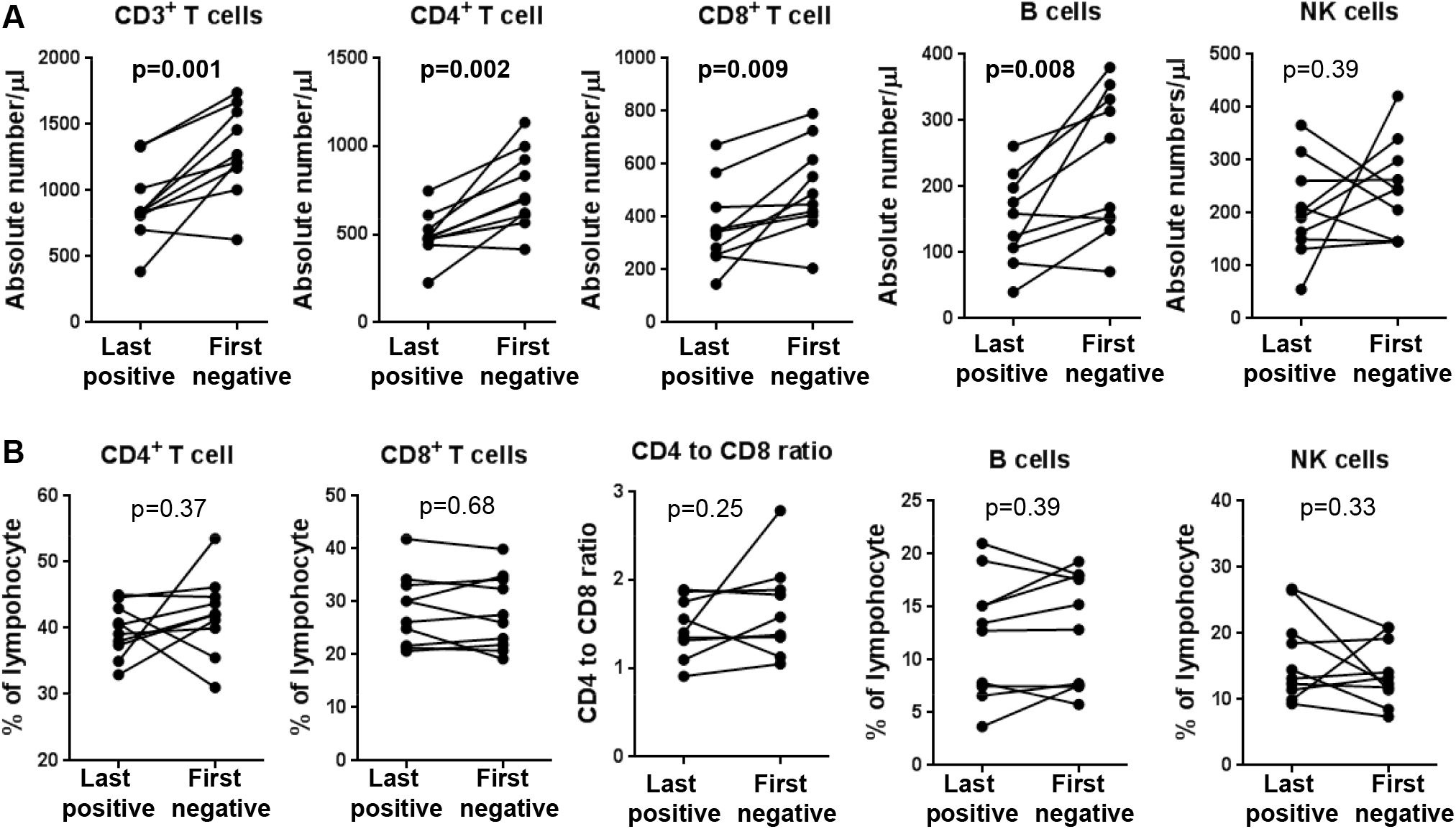
The alterations of absolute numbers (A) and relative frequencies of lymphocyte subpopulations (B) in peripheral blood of PP patients after they turned negative for SARS-CoV-2 RNA detection. *P* < 0.05 was considered statistically significant (in bold).

Finally, we were able to extract paired results of lymphocyte subpopulations for 10 patients on admission (last positive), and on the first day they tested negative for viral RNA (first negative). These patients demonstrated markedly increased CD3^+^ T cells (p=0.001), CD4^+^ T cells (p=0.002), CD8^+^ T cells (p=0.009), and B cells (p=0.008) after turned negative for SARS-CoV-2. Together, these results indicated that the abnormalities in adaptive immune cells, but not symptoms and laboratory indicators, were associated with SARS-CoV-2 viral RNA detection in non-severe COVID-19 patients.

## Discussion

This retrospective investigation was designed to examine immunological characteristics of non-severe COVID-19 patients with persistent viral presence. We reported here that despite their alleviated symptoms and much improved laboratory findings, these patients demonstrated significantly lower numbers of T cells and B cells than healthy controls, and than those turned negative for viral RNA.

37 non-severe COVID-19 patients with persistent viral presence were included in this study and were allocated to the PP group. Multiple symptoms, including fever, dry cough, dyspnea, expectoration, diarrhea, pharyngalgia, hemoptysis and weep tears were recorded at disease onset (Table 1), and most of these patients were abnormal in radiographic examination (data not shown). Upon treatment in mobile cabin hospital and transferred to our hospital, they went almost asymptomatic, and had much improved laboratory findings, as showed in Table 2 and compared with those in the PA group. However, persistent SARS-CoV-2 presence were evident in all these patients.

The presence of SARS-CoV-2 has been the golden standard for both diagnosis and disease management of COVID-19. In fact, two consecutively negative results for viral RNA is required for patients to be discharged from hospitals [5]. Nasopharyngeal swabs were frequently used for detecting viral RNA by RT-PCR because these samples are easily accessible. However, some limitations were noticed. First, the kinetics of SARS-CoV-2 shedding was different from that of SARS-CoV and MERS-CoV. RNA copies of SARS-CoV-2 were very high in nasopharyngeal swab during the first week of symptoms, with peak on day 4 post-onset, whereas the peak value appeared until 7-10 days post-onset with much lower RNA copies during SARS-CoV and MERS-CoV infection [6,18-20]. Second, the presence of virus RNA in lower respiratory tract (sputum or BALs), stool, and blood samples were reported, and the kinetics of virus shedding in these sites were distinct from that in throat [8-11]. Third, sampling error and the technical limitations of RT-PCR sometimes led to a false testing result [21]. With these limitations, it is not surprising that some patients who tested negative in two consecutively RT-PCR tests and were discharged from hospital had positive results 5 to 13 days later [21,22]. In together, these notions posed a great challenge to discharge management for COVID-19 patients, especially for non-severe cases having obtained clinical cure.

Since the presence of viral RNA might come from fragments of dead virus, isolating live SARS-CoV-2 is therefore useful in determining viral infectivity [6]. However, this method is required to be performed in a biological safety level 3 (BSL-3) laboratory, which limited its application in clinical practice for discharge management. Indicators from the immune system are promising candidates in this regard. Detection for virus-specific IgM and IgG has been widely used in hepatitis and other virus infectious diseases for helping the diagnosis of viral infection, as well as for evaluating disease status and prognosis [23]. It was reported that SARS-CoV-specific IgM and IgG was generated 3-6 days and 8-14 days post infection, respectively [24]. In fact, detection for virus-specific IgM and IgG were recently included in the latest version of the guideline of SARS-CoV-2 (Trial Version 7 of the Chinese National Health Commission), for assisting the diagnosis of SARS-CoV-2 infection [25]. However, antigen selection and assay sensitivity may cause both false positive and false negative results [26]. Thus, its efficacy in diagnosis and discharge management remained to be tested by large clinical investigations. The production of both antibody isotypes requires the cooperation between virus-specific T cells and B cells. Therefore, alterations of these adaptive immune cells might precede the changes of antibodies and could be useful for discharge management.

Lymphopenia was observed at illness onset in 72.8% of non-severe COVID-19 patients (the PA group) in our study, which is similar to those reported by Zhang et al [15] (75.4%), Mo et al [17] (73.5%), Wang et al [27] (70.3%), and Guan et al [2] (83.2%), suggesting the involvement of lymphocytes in the early phase of SARS-CoV-2 infection. Furthermore, lymphocyte count was reported to be correlated with disease severity. Significant higher numbers of lymphocytes were found in survivors versus non-survivors [4], as well as critically ill versus severe [13,14], and severe versus non-severe cases [15,16]. We focused on non-severe patients with persistent viral presence, and found that the PP group had markedly higher lymphocyte count (1.5 [1.3-1.8] vs 0.9 [0.7-1.3]; p<0.001) than the PA group, and were comparable to healthy subjects. This finding, together with alleviated symptoms and improvements of other laboratory findings, indicated that PP patients might be in the process of recovery, albeit their viral RNA were still tested positive. However, other parameters are required to determine if they were fully recovered. We therefore examined lymphocyte subsets and found that PPP patients had significantly lower numbers of CD3^+^ T cells (p=0.001), CD4^+^ T cells (p=0.005), CD8^+^ T cells (p=0.003), and B cells (p=0.005) than PPN patients (Fig 1A and 1B). When compared with healthy subjects, PPP patients again exhibited much less CD3^+^ T cells (p=0.044), CD4^+^ T cells (p=0.034), and B cells (p=0.02) (Fig 2C and 2D). Most strikingly, 10 PPN patients showed markedly increased CD3^+^ T cells (p=0.001), CD4^+^ T cells (p=0.002), CD8^+^ T cells (p=0.009), and B cells (p=0.008) after they turned negative for SARS-CoV-2. Together, these results suggest that measurement of these lymphocyte subpopulations could be used to distinguish non-severe patients with persistent viral presence from healthy subjects and those turned negative, and thus have clinical relevance for discharge management.

T cells and B cells are the two most important lymphocytes in fighting against viral infection. CD8^+^ T cells are particularly efficient in clearing virus-infected cells, after receiving help from CD4^+^ T cells [28]. The latter can induce the activation and differentiation of cognate B cells, and subsequently promote the production of virus-specific antibodies, including neutralizing antibodies [29]. In turn, neutralizing antibodies are able to mediate antibody-dependent cell-mediated cytotoxicity to kill virus-infected cells, and to block the entrance of extracellular virus [30]. Therefore, it’s not surprising that changes in these cells could reflect the viral presence. Accordingly, T cell subsets were reported to be profoundly affected in severe cases with SARS-CoV-2 infection [16]. However, we could not determine from our data and the current knowledge whether SARS-CoV-2 can directly infect these lymphocytes, or indirectly caused these alterations. We did not find any difference in NK cells between the PPP group and healthy subjects, in terms of both absolute numbers and relative frequency (Fig 2A and 2D). Instead, NK cells were even higher in the PPP group than in the PPN group (p=0.02, Fig 1A). As an innate immune cells, NK cells is among the first cell types to combat virus infection [31]. However, PP patients in our study were likely to be in the late phase of SARS-CoV-2 infection, during which the role of NK cells remained to be defined. Several limitations to the present study warrant mention. First, this retrospective study was conducted in a single hospital, which may result in selection bias. Our conclusion could be further strengthened by a multicenter, prospective study in a randomized setting. Second, only 37 non-severe COVID-19 patients with persistent viral presence were included in this investigation, interpretation of our findings might be limited by the sample size. Third, these patients were transferred to our hospital, we do not have their laboratory results and lymphocyte measurements at disease onset, we therefore randomly selected age- and sex-matched PA patients for comparison. Fourth, quantitative viral RNA detection and isolation of live virus were not performed due to limited resources in our hospital, which prevent us from building connections between lymphocyte subpopulations and these parameters.

Despite these limitations, the present study, to the best of our knowledge, is the first investigation to examine changes of lymphocyte subpopulations in non-severe COVID-19 patients with persistent viral presence. We found that CD4^+^ T cells, CD8^+^ T cells, and B cells were markedly decreased in these patients. Our findings suggest that monitoring lymphocyte subpopulations could be clinical meaningful in discharge management for non-severe COVID-19 patients with persistent viral presence.

## Data Availability

After publication, the data will be made available to others on reasonable requests to the corresponding author. A proposal with detailed description of study objectives and statistical analysis plan will be needed for evaluation of the reasonability of requests. Additional materials might also be required during the process of evaluation. Deidentified participant data will be provided after approval from the corresponding author and Zhongnan Hospital of Wuhan University.

## Funding

This work was supported by the National Natural Science Foundation of China (2017NSFC81670825 to Y.H. and 2020NSFC31970865 to J.H.), Natural Science Foundation of Hubei Province (2017CFB512 to B.L.), and Science and Technology Innovation Cultivation Foundation of Zhongnan Hospital of Wuhan University (ZNPY2018107 to B.L.).

### Acknowledgement

We thank all patients and their families involved in the study, and all the healthcare professionals from Zhongnan Hospital of Wuhan university.

## Abbreviations

COVID-19: Coronavirus disease 2019
SARS-CoV-2: Severe acute respiratory syndrome coronavirus 2
HC: Healthy controls

